# Real-world effectiveness of mpox (monkeypox) vaccines: a systematic review

**DOI:** 10.1101/2023.03.16.23287333

**Authors:** Mingda Xu, Caifen Liu, Zhanwei Du, Yuan Bai, Zhen Wang, Chao Gao

## Abstract

Understanding the effectiveness of smallpox vaccines against mpox in humans can inform public health preparedness for mpox prophylaxis during the 2022 outbreak. However, the efficacy of current vaccines against mpox has been inferred from animal and immunogenicity studies and has barely been demonstrated in human clinical trials. Thus, we conducted a systematic literature review to estimate the real-world effectiveness of mpox vaccines. The study showed a pooled estimate of 0.89 (95% CI: 0.86-0.91) for the third-generation smallpox vaccine (MVA-BN) effectiveness during the 2022 outbreak. The results showed that the incidence of mpox was significantly lower in the vaccinated group than in the unvaccinated group. Considering the potential mutations, side effects, and breakthrough infections, it is recommended to develop a next-generation vaccine that is more effective and safer for future mpox outbreaks.

## Main Text

Mpox (monkeypox) virus is a DNA virus in the orthopoxvirus genus, which was first identified among humans in the Democratic Republic of the Congo in 1970 [1]. In May 2022, a series of mpox cases were identified in non-endemic countries and has rapidly spread worldwide. On July 23, 2022, the World Health Organization declared the mpox outbreak a Public Health Emergency of International Concern (PHEIC) [2]. As of 1 February 2023, 85,531 cases and 91 deaths had been reported in over 110 locations [3]. The urgent need to mitigate mpox transmission, especially for populations at risk, has surged the accelerating development of mpox vaccines and treatments. However, clinical efficacy data for mpox vaccines are currently unavailable. The 1st-generation vaccine (e.g., Dryvax) is no longer available since the cessation of smallpox vaccination after eradication in 1980. Then, the 2nd-generation vaccine (ACAM2000) was approved by US FDA against orthopoxvirus in August 2007 [4]. The 3rd-generation smallpox vaccines like MVA-BN (also known as JYNNEOS and IMVANEX) [5] are expected to be the critical factors that can significantly convert the mpox prevention strategies. Understanding the effectiveness among humans can inform public health for future preparedness of mpox prophylaxis. This study aims to review the real-world effectiveness of mpox vaccines.

We identified 236 articles by searching PubMed, 120 by Web of Science, and 6 studies from expert recommendation and retrieved 179 related preprints from medRxiv and bioRxiv as of 25 January 2023. We identified 486 distinct studies for screening. 8 studies reported the results of vaccine effectiveness that met the inclusion requirements for this review, as shown in **Figure S1**. The effectiveness results for 1st-generation (Dryvax and unknown) and 3rd-generation (MVA-BN) were included. **Table 1** summarises the articles included in this study.

**Table 1.**
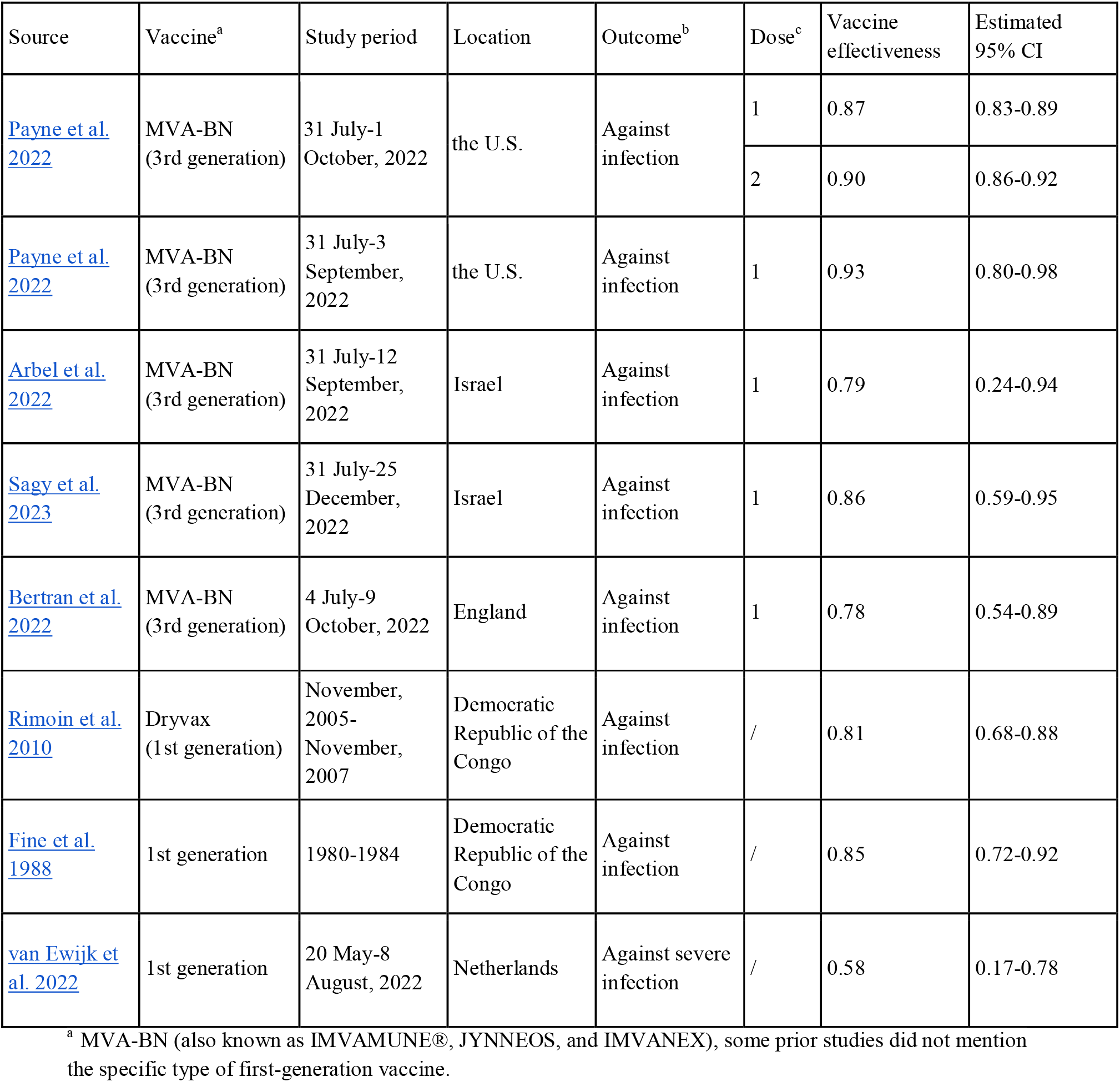
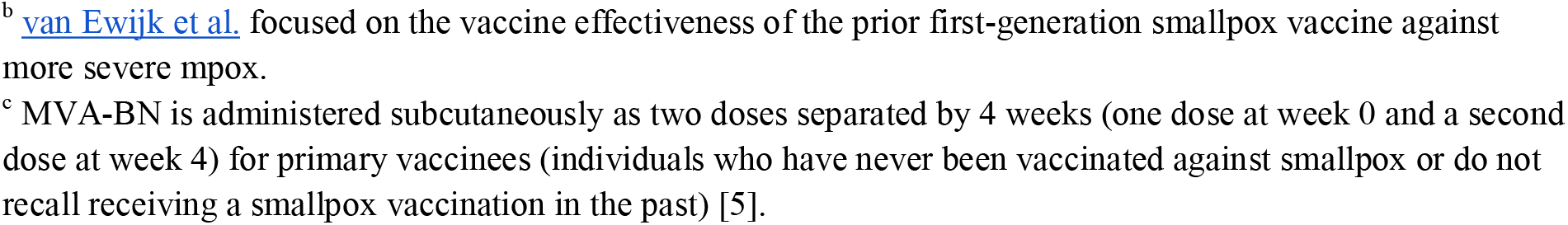
Summary of the effectiveness results for the mpox vaccines.

| Source | Vaccine<sup>a</sup> | Study period | Location | Outcome<sup>b</sup> | Dose<sup>c</sup> | Vaccine effectiveness | Estimated 95% CI |
| --- | --- | --- | --- | --- | --- | --- | --- |
| Payne et al. 2022 | MVA-BN (3rd generation) | 31 July-1 October, 2022 | the U.S. | Against infection | 1 | 0.87 | 0.83-0.89 |
|  |  |  |  |  | 2 | 0.90 | 0.86-0.92 |
| Payne et al. 2022 | MVA-BN (3rd generation) | 31 July-3 September, 2022 | the U.S. | Against infection | 1 | 0.93 | 0.80-0.98 |
| Arbel et al. 2022 | MVA-BN (3rd generation) | 31 July-12 September, 2022 | Israel | Against infection | 1 | 0.79 | 0.24-0.94 |
| Sagy et al. 2023 | MVA-BN (3rd generation) | 31 July-25 December, 2022 | Israel | Against infection | 1 | 0.86 | 0.59-0.95 |
| Bertran et al. 2022 | MVA-BN (3rd generation) | 4 July-9 October, 2022 | England | Against infection | 1 | 0.78 | 0.54-0.89 |
| Rimoin et al. 2010 | Dryvax (1st generation) | November, 2005- November, 2007 | Democratic Republic of the Congo | Against infection | / | 0.81 | 0.68-0.88 |
| Fine et al. 1988 | 1st generation | 1980-1984 | Democratic Republic of the Congo | Against infection | / | 0.85 | 0.72-0.92 |
| van Ewijk et al. 2022 | 1st generation | 20 May-8 August, 2022 | Netherlands | Against severe infection | / | 0.58 | 0.17-0.78 |

<sup>a</sup> MVA-BN (also known as IMVAMUNE®, JYNNEOS, and IMVANEX), some prior studies did not mention the specific type of first-generation vaccine.
<sup>b</sup> van Ewijk et al. focused on the vaccine effectiveness of the prior first-generation smallpox vaccine against more severe mpox.
<sup>c</sup> MVA-BN is administered subcutaneously as two doses separated by 4 weeks (one dose at week 0 and a second dose at week 4) for primary vaccinees (individuals who have never been vaccinated against smallpox or do not recall receiving a smallpox vaccination in the past) [5].

The results showed that the incidence of mpox was significantly lower in the vaccinated group than in the unvaccinated group. Past epidemiological data from African endemic areas showed an almost 20-fold increase of mpox cases after 1st-generation smallpox vaccination stopped, indicating an estimated 80-85% vaccine effectiveness. The effectiveness of the prior 1st-generation smallpox vaccine against more severe mpox was 0.58 (95% CI: 0.17-0.78) in the Netherlands. We further estimated the pooled estimates of 0.89 (95% CI: 0.86-0.91) for the 3rd-generation mpox vaccine effectiveness during the 2022 mpox outbreak (**Figure S2**).

Our study has some limitations, including significant variations in the included studies. The reported results are real-world vaccine effectiveness against mpox, which may vary over studies due to factors (e.g., estimation methods, population under observation).

In summary, the results from the included studies demonstrated the encouraging effectiveness of the mpox vaccines being widely used. The development of effective vaccines and treatments should be a priority to protect people’s health. Vaccines, antivirals, and other treatments should be implemented promptly, and monitoring of the virus must be ongoing. Additionally, environmental and behavioural interventions such as improved sanitation, vector surveillance and public health measures such as travel restrictions and case isolation are essential components of the response.

## Supporting information

Supplemental File for Manuscript

## Data Availability

All data produced in the present study are available upon reasonable request to the authors

## Acknowledgements

We acknowledge the financial support from the Key Program for International Science and Technology Cooperation Projects of China (No. 2022YFE0112300), National Natural Science Foundation of China (Nos. 61976181, 62261136549, U22B2036), Key Technology Research and Development Program of Science and Technology - Scientific and Technological Innovation Team of Shaanxi Province (No. 2020TD-013).

## Author contributions

M.X. and C.L. did the literature search and created the table and figure. M.X. and Z.D. conducted analyses, interpreted results, wrote and revised the manuscript. Y.B., Z.W. and C.G. interpreted results and revised the manuscript.

## Data Availability

The data underlying this article and these programs will be shared on reasonable request with the corresponding author.

