## Supplemental File for Manuscript for "Real-world effectiveness of mpox (monkeypox) vaccines: a systematic review"

**Supplementary material**

### **Methods**

#### **Search Strategy and Selection Criteria**

All searches were conducted on 25 January, 2023 in PubMed and Web of Science for published articles and medRxiv for preprints. We also included 6 reports from expert recommendations.

The following search terms were used:

- 1) PubMed and Web of Science: (monkeypox OR mpox) AND (vaccine OR vaccination) AND (efficacy OR effectiveness);
- 2) medRxiv and bioRxiv: ("mpox" or "monkeypox") and ("vaccine" or "vaccination") and ("effectiveness" or "efficacy").

After screening the abstract and full text, we included the studies that met the following inclusion criteria:

- 1) The studies reported on the use of smallpox vaccines against mpox in real-world humans;
- 2) The vaccine efficacy/effectiveness results were published either in published articles or in the preprints from medRxiv;
- 3) The studies published the results of vaccine efficacy/effectiveness against mpox infection at least 1 dose;
- 4) The results of the real-world were positive and warranted future development.

#### **Data Extraction**

All data were extracted independently and converted to a standardized form by 2 authors (M.X. and C.L.). Disagreements in study inclusion and data extraction were resolved by a third author (Z.D.). The primary results were the real-world effectiveness of mpox vaccine. For effectiveness, the value combined with the confidence interval, and the infection it was against were collected. The risks of infection in the unvaccinated group and the vaccinated group were also collected and used to calculate the effectiveness in case it was not provided in the study. We collected the estimations on real-world vaccine effectiveness and efficacy, along with the corresponding 95% confidence interval (CI). The vaccine effectiveness was calculated as 1– relative risk (RR), with the relative risk estimated by the maximum likelihood method in R package *epitools*. The information related to the settings of observation, including the study period, locations, and type of vaccine, were also extracted from each eligible study [[1–8]](https://paperpile.com/c/MH3BeE/6dtN+fqr6+zWKm+dKap+DAs4+vZCd+PwW3+jNdY).

#### **Statistical Analysis**

Following previous studies [[9,10]](https://paperpile.com/c/MH3BeE/jgIj+wwp6), study heterogeneity was categorized into three levels based on the *I^2^* index: *I^2^* > 75% (high heterogeneity), *I^2^* = 25-75% (average heterogeneity), and *I^2^* < 25% (low heterogeneity), and a meta-analysis was performed using the random effects model (Hartung-Knapp). We estimated the pooled estimates of the vaccine effectiveness for studies that provided the 95% CI, using the function metagen of the R package *meta*. All analyses were conducted in R version 4.2.2.

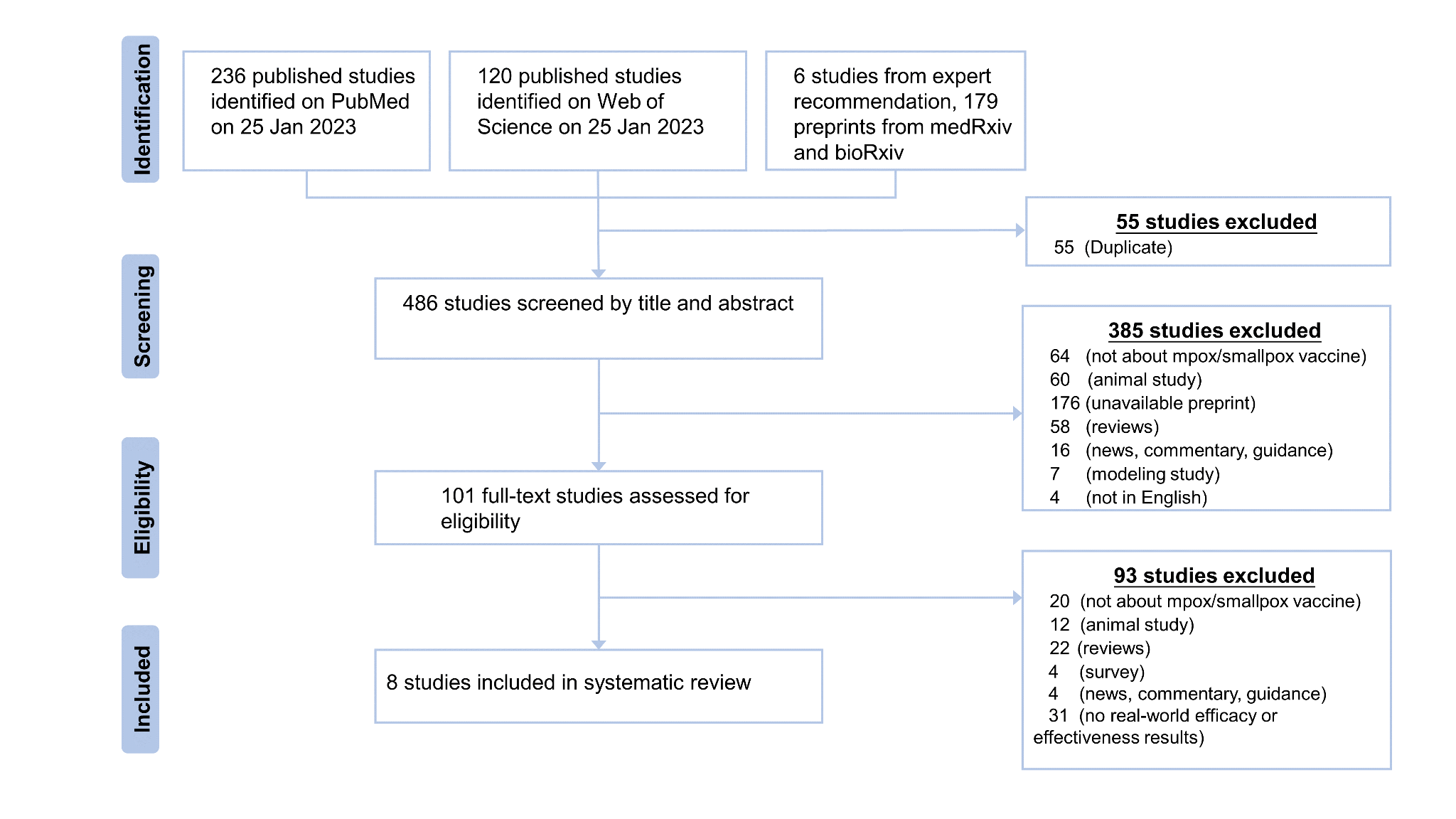
**Figure S1. PRISMA (Preferred Reporting Items for Systematic Reviews and Meta-Analyses) flow diagram for searching and selecting studies that reported real-world mpox vaccine effectiveness and efficacy.**

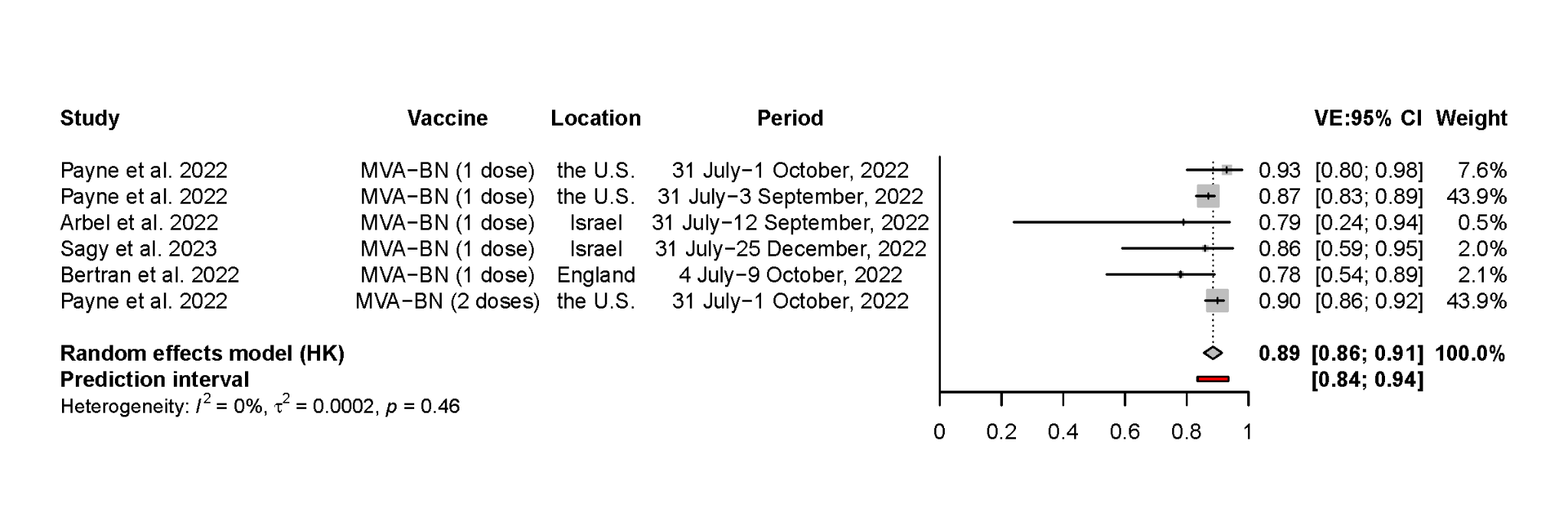
**Figure S2. Pooled estimates of the 3rd-generation smallpox vaccine’s effectiveness against mpox from 5 included studies.**
